# Left Hemisphere Bias of NIH Stroke Scale is Most Severe for Middle Cerebral Artery Strokes

**DOI:** 10.1101/2021.12.06.21267370

**Authors:** Emilia Vitti, Ganghyun Kim, Melissa D. Stockbridge, Argye E. Hillis, Andreia V. Faria

**Affiliations:** Department of Neurology, School of Medicine, Johns Hopkins University, Baltimore, MD, USA; Department of Neuroscience, Johns Hopkins University, Baltimore, MD, USA; Department of Physical Medicine & Rehabilitation, and Department of Cognitive Science, Johns Hopkins University, Baltimore, MD, USA; Department of Radiology, School of Medicine, Johns Hopkins University, Baltimore, MD, USA

## Abstract

**Background and Aim:** NIHSS score is higher for left versus right hemisphere strokes of equal volumes. However, differences in each vascular territory have not been evaluated yet. We hypothesized that left versus right differences are driven by the middle cerebral artery (MCA) territory, and there is no difference between hemispheres for other vascular territories.

**Methods:** This study is based on data from 802 patients with evidence of acute or early subacute ischemic stroke. These patients had infarct restricted to one major arterial territory (MCA, n=437; PCA, n=209; ACA, n=21; vertebrobasilar, n=46) and received NIHSS and MRI at hospital admission. We examined differences in patients with left or right strokes regarding to lesion volume, NIHSS, and other covariates (age, sex, race). We used linear models to test the effects of these covariates on NIHSS. We looked at the whole sample as well as in the sample stratified by NIHSS (<=5 or >5) and by lesion location (MCA or PCA).

**Results:** Patients with left MCA strokes had significantly higher NIHSS than those with right strokes. Only patients with MCA strokes showed NIHSS score affected by the hemisphere when controlling for stroke volume and patient’s age. This difference was driven by the more severe strokes (NIHSS>5). In addition, stroke volume and patient’s age significantly correlated with NIHSS.

**Conclusion:** Right MCA infarcts are larger than left MCA infarcts associated with a given NIHSS score, after accounting for other significant associations, such as patient’s age. It is important to consider this systematic bias in the NIHSS when using the score for inclusion criteria for treatment or trials. Patients with right MCA stroke may be under-treated and left with disabling deficits that are not captured by the NIHSS.

## 1 Introduction

The National Institutes of Health Stroke Scale (NIHSS) is a valid and reliable tool most frequently used for clinically evaluating acute stroke^1–4^. The NIHSS is associated with severity, long-term functional outcomes^5–7^, infarct size, lesion location^1^, and angiographic findings^8–10^. Scoring the NIHSS consists of broad categories associated with stroke signs and symptoms (e.g., level of consciousness, motor performance, language, speech, neglect, etc.). Distinct clinical features or stroke syndromes can be appreciated depending on the specific vascular territory affected (i.e., anterior cerebral artery (ACA), posterior cerebral artery (PCA), middle cerebral artery (MCA)).

The NIHSS is designed to represent left and right cortical and motor function equally; however, there are more opportunities to award points for left hemisphere dysfunction than right^8, 11, 12^. This is likely because up to 7 points are directly related to language deficits, and these deficits typically are associated with left MCA stroke only, particularly among right-handed people. Points attributable to left MCA cortical strokes are awarded across three categories: orientation questions, 2; following commands, 2; and specific language tasks to determine signs of aphasia, 3 (e.g., picture description, confrontation naming, sentence reading), in addition to sensory and motor. In contrast, points attributable to right MCA cortical stroke are awarded in only one category: neglect, 2, other than sensory and motor (Supplementary Table 4). Thus, even if the stroke volume is equal, NIHSS scores are often higher for left vs right hemisphere stroke^8, 11^.

Differences in NIHSS relative to specific vascular territories have not yet been evaluated. Deficits associated with bilateral PCA and ACA stroke are likely symmetrically represented in the NIHSS scale, because language is largely specific to the left MCA territory versus right. However, left ACA and PCA stroke can cause aphasia (e.g., transcortical motor aphasia, optic aphasia; see^13^). Hemispatial neglect can be caused by ACA, MCA, or PCA stroke. While neglect is more noticeable after right hemisphere stroke, right neglect after left hemisphere stroke may be almost as common^14^. Thus, there are clear reasons to suspect that hemispheric bias in the NIHSS may be specific to MCA territory strokes. We hypothesized that right MCA strokes have larger infarct volumes than left MCA ischemic strokes, in groups with similar NIHSS, but there will be no difference between hemispheres for other vascular territories. Similarly, we hypothesized that after controlling for stroke volume and other covariates, the side of MCA infarcts, but not of infarcts in other territories, significantly affects the NIHSS.

## 2 Methods

This study included MRIs of patients admitted to the Comprehensive Stroke Center at Johns Hopkins Hospital with the clinical diagnosis of ischemic stroke, between 2009 and 2019 (Flowchart for data inclusion in Figure 1). It utilizes data from an anonymized dataset, created under waiver of informed consent (IRB00228775). We have complied with all relevant ethical regulations and the guidelines of the Johns Hopkins Institutional Review Board, that approved the present study (IRB00290649).

**Figure 1.**
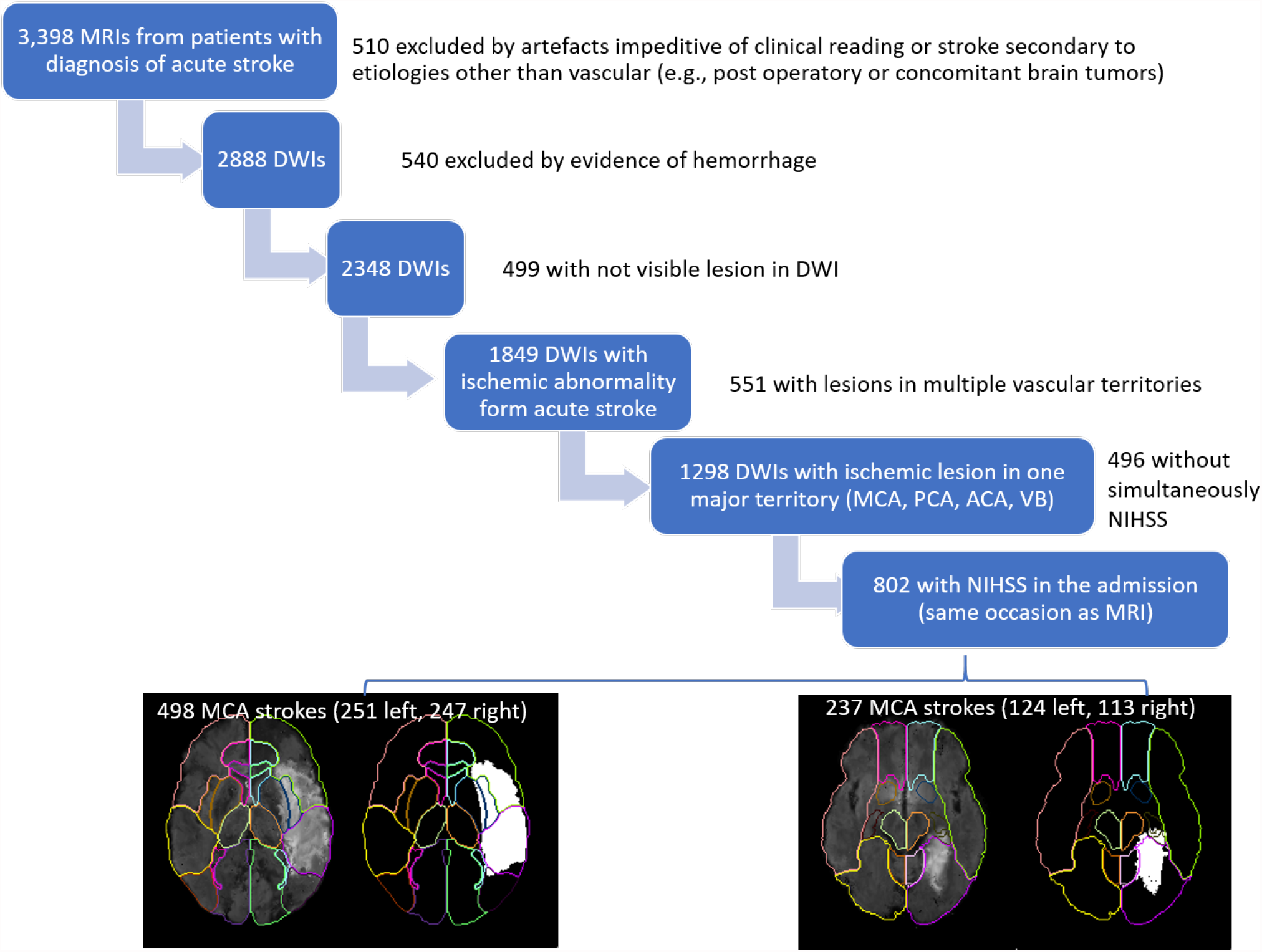
Flowchart of data inclusion

From the 2,888 DWIs quality-controlled for clinical analysis, 1,849 DWIs showed lesions classified by a neuroradiologist as result of acute or early subacute ischemic stroke, with no evidence of hemorrhage. From those, we included 802 individuals who had NIHSS recorded at admission, in the same occasion as the MRI, and the infarct lesion constrained to a single vascular territory (MCA, PCA, ACA, vertebrobasilar). The present study focuses on the largest groups of lesions, affecting the MCA (n=498) and PCA (n=237) territories. The summary of demographics and lesion profiles is in Table 1. The lesion core was defined in DWI, in combination with the Apparent Diffusion Coefficient maps (ADC) by two experienced evaluators and was revised by a neuroradiologist until reaching a final decision by consensus. Further details are in our previous publication^15^.

**Table 1.** Demographic and lesion characteristics. Continuous variables are showed as mean (standard deviation) [minimun - maximun]. * indicates significant difference at p<0.05

|  | All |  |  | MCA |  |  | PCA |  |  |
| --- | --- | --- | --- | --- | --- | --- | --- | --- | --- |
|  | Total (N=802) | Left (N=420) | Right (N=382) | Total (N=498) | Left (N=251) | Right (N=247) | Total (N=237) | Left (N=124) | Right (N=113) |
| NIHSS | 5.5 (5.9) [0-31] | 5.8 (6.4) [0-31] | 5.1 (5.4) [0-24] | 6.9 (6.7) [0-31] | 7.5* (7.2) [0-31] | 6.3* (6.1) [0-24] | 3.0 (2.8) [0-19] | 3.1 (3.1) [0-19] | 2.9 (2.4) [0-11] |
| lesion volume (cc) | 23.8 (52.2)<br>[0.028-403.4] | 22.8 (513)<br>[0.028-383.4] | 25.02 (53.2)<br>[0.042-403.46] | 34.66 (62.9)<br>[0.06-403.46] | 33.93 (63.8)<br>[0.084-383.4] | 35.41 (63.1)<br>[0.06-403.46] | 6.8 (14.9)<br>[0.028-131.6] | 7.2 (16.9)<br>[0.028-131.6] | 6.4 (12.6)<br>[0.042-73.7] |
| Age | 62 (13) | 61 (13) | 62 (14) | 62 (14) | 62 (13) | 62 (15) | 62 (12) | 60 (13)* | 63 (12)* |
| Sex |  |  |  |  |  |  |  |  |  |
| Female | 370 (46.1%) | 191 (45.5%) | 179 (46.9%) | 233 (46.8%) | 118 (47.0%) | 115 (46.6%) | 101 (42.6%) | 50 (40.3%) | 51 (45.1%) |
| Male | 432 (53.9%) | 229 (54.5%) | 203 (53.1%) | 265 (53.2%) | 133 (53.0%) | 132 (53.4%) | 136 (57.4%) | 74 (59.7%) | 62 (54.9%) |
| Race |  |  |  |  |  |  |  |  |  |
| African American | 485 (60.5%) | 249 (59.3%) | 236 (61.8%) | 279 (56.0%) | 130 (51.8%) | 149 (60.3%) | 165 (69.6%) | 92 (74.2%) | 73 (64.6%) |
| Caucasian | 276 (34.4%) | 148 (35.2%) | 128 (33.5%) | 190 (38.2%) | 104 (41.4%) | 86 (34.8%) | 65 (27.4%) | 29 (23.4%) | 36 (31.9%) |
| Other / Unknown | 41 (5.1%) | 23 (5.5%) | 18 (4.7%) | 29 (5.8%) | 17 (6.8%) | 12 (4.9%) | 7 (3.0%) | 3 (2.4%) | 4 (3.5%) |
| Vascular Territory |  |  |  |  |  |  |  |  |  |
| MCA | 498 (62.1%) | 251 (59.8%) | 247 (64.7%) |  |  |  |  |  |  |
| PCA | 237 (29.6%) | 124 (29.5%) | 113 (29.6%) |  |  |  |  |  |  |
| Vertebrobasilar | 46 (5.7%) | 27 (6.4%) | 19 (5.0%) |  |  |  |  |  |  |
| ACA | 21 (2.6%) | 18 (4.3%) | 3 (0.8%) |  |  |  |  |  |  |

In an initial exploratory analysis, we examined differences in groups of patients with left or right stroke regarding to lesion volume, NIHSS and other covariates (age, sex, race). We used t-test for continuous variables and chi-square tests for categorical variables. We then used generalized linear models to test the effects of lesion side, volume, patient’s age, sex, and race on NIHSS. We used Akaike information criterion (AIC) to evaluate the impact of covariates or of their interactions in the models. We looked at the whole sample as well as in the sample stratified by NIHSS (<=5 or >5) and by lesion location (MCA or PCA). The statistical analysis was performed with R.

## 3 Results

As shown in Table 1, there were no significant differences between left and right strokes in terms of patient’s age, sex, and race. The distribution of these variables was similar in left and right strokes. Patients with left MCA strokes had significantly higher NIHSS than those with right MCA strokes. This difference was not significant in patients with PCA strokes. In addition, left MCA strokes tended to be larger than the right counterparts (although not significantly at p<0.05). This tendency persisted after NIHSS stratification (<=5 or >5) as shown in Table 2. This left hemisphere bias is illustrated in Figure 2.

**Figure 2.**
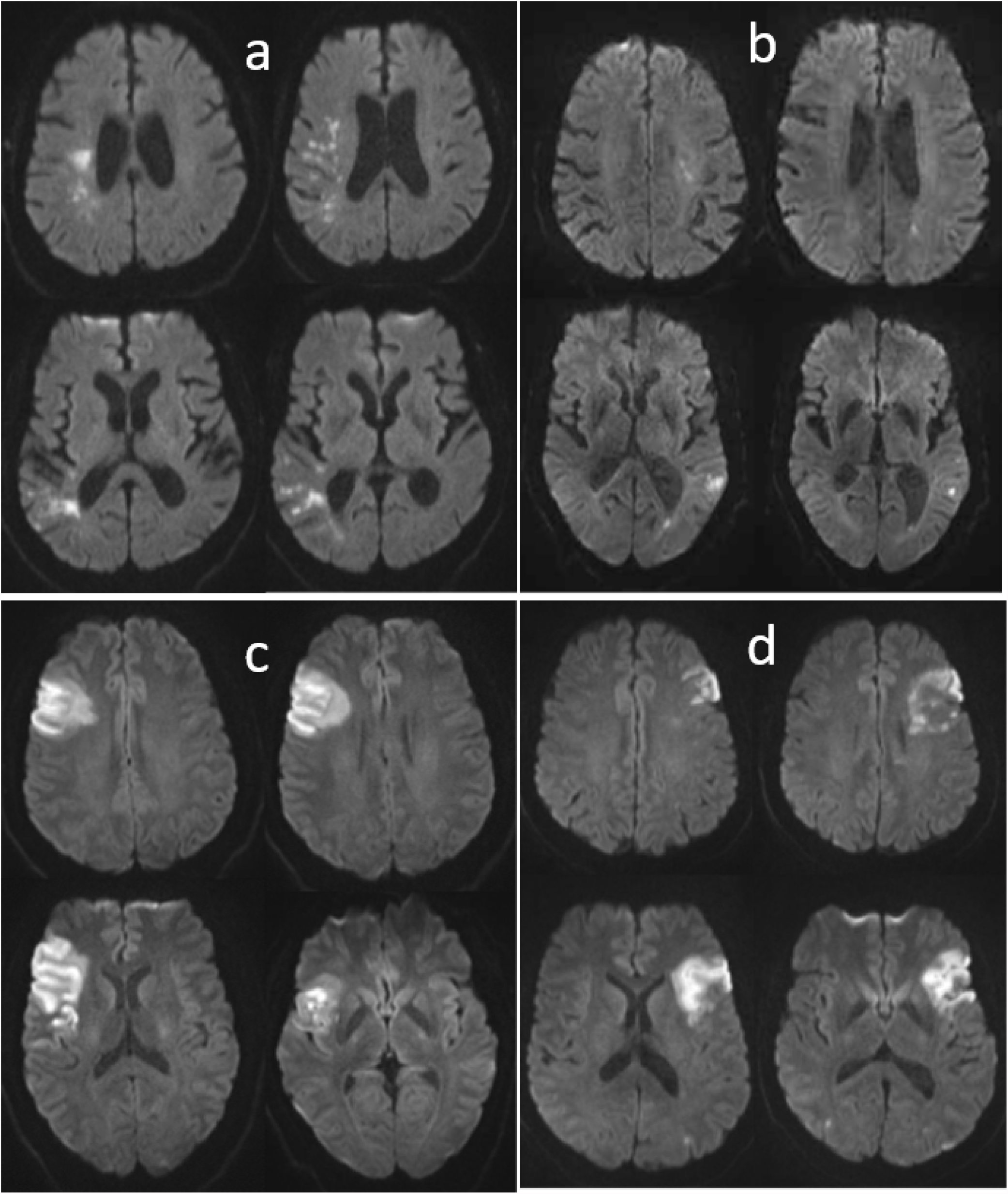
Illustrative cases of patients with right (a and c) and left (b and d) strokes, with similar NIHSS and very different infarct volume (a and b); or similar infarct volumes and very different NIHSS (c and d). a) 76 years-old man, NIHSS=4, infarct volume of 11.9 cc; b) 84 years-old man, NIHSS=5, infarct volume of 2.2 cc; d) 34 years-old man, NIHSS=6, infarct volume of 39 cc; b) 57 years-old man, NIHSS=11, infarct volume of 31 cc.

**Table 2.**
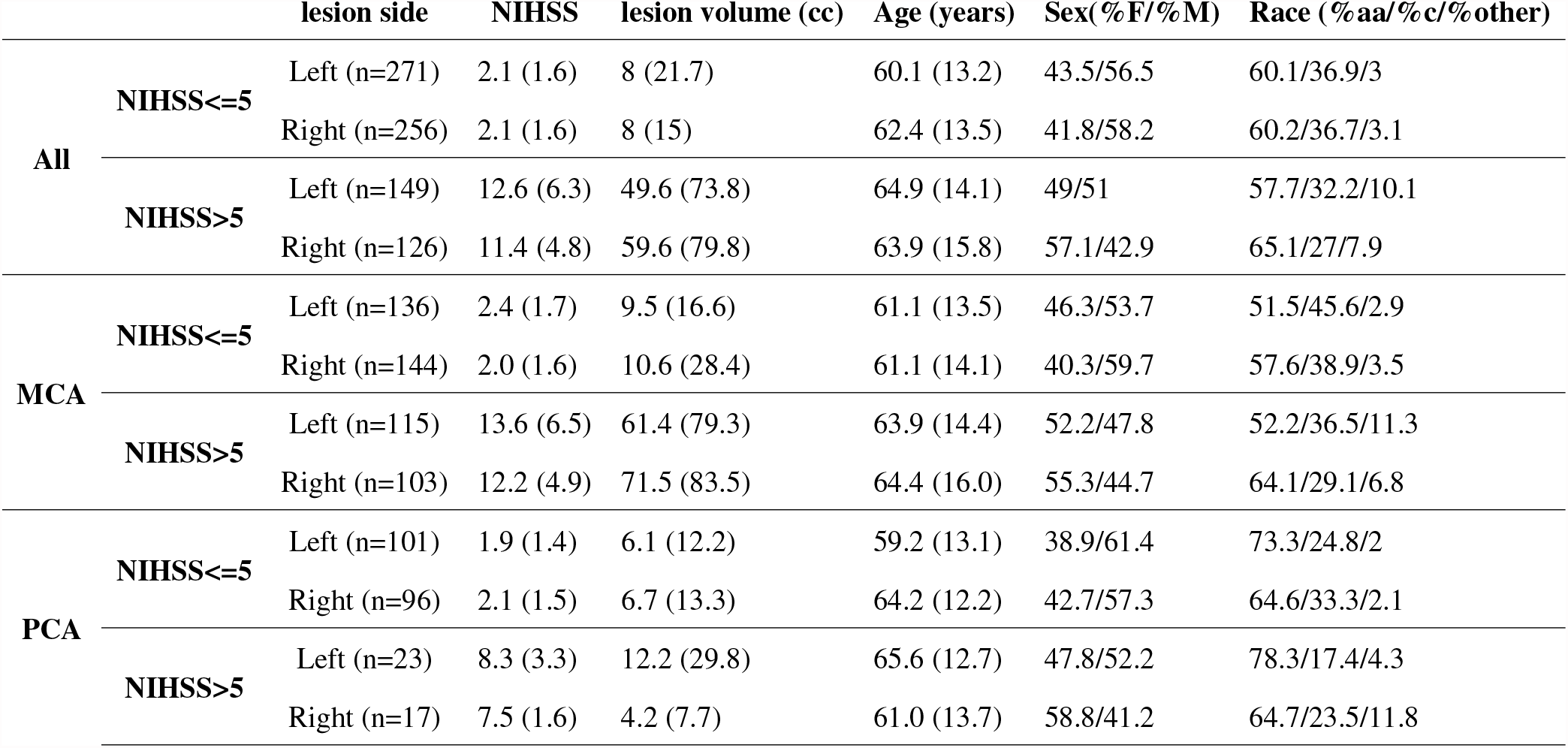
Demographic and lesion characteristics in samples stratified by the vascular territory affected and by NIHSS. F=female, M=male, aa=African Americans, c=caucasians, others include others and unknown races. Continuous variables are represented as mean (standard deviation)

The initial linear models to assess the effects of covariates in NIHSS included stroke side, volume, patient’s age, sex, race and interactions between stroke side and volume. As the effect of race was not significant in any model, race was further excluded from the analysis. Patient’s sex and interactions between stroke side and volume were marginally associated to NIHSS (p-value for sex =0.034; p-value for the interaction between side and volume = 0.03), only when considering the whole sample. The models with or without these covariates were equivalent (both showed AIC = 4753). Therefore, sex and interaction between stroke volume and side were also excluded from further models. In summary, the final models used age, lesion side and volume as predictors. They revealed that in patients with MCA strokes, and not in those with PCA strokes, NIHSS score is affected by the infarct side (p-value for infarct side = 0.00491) even after controlling for stroke volume and patient’s age, as shown Table 3 and Supplementary Table 5. This effect was driven by the more severe strokes (NIHSS>5). In addition, stroke volume and patient’s age significantly correlated with NIHSS.

**Table 3.** P-values for the generalized linear models and covariates of the models to predict the NIHSS.

|  | All |  |  | MCA |  |  | PCA |  |  |
| --- | --- | --- | --- | --- | --- | --- | --- | --- | --- |
|  | Total | NIHSS<=5 | NIHSS>5 | Total | NIHSS<=5 | NIHSS>5 | Total | NIHSS<=5 | NIHSS>5 |
| sample | 802 | 527 | 275 | 498 | 280 | 218 | 237 | 197 | 40 |
| intercept | 0.7071 | 7.12E-07 | 3.81E-07 | 0.7844 | 0.01365 | 1.54E-06 | 0.0167 | 2.31E-06 | 0.0532 |
| lesion volume | <2.2e-16 | 0.00167 | 2.00E-16 | 2.00E-16 | 0.00384 | 7.67E-15 | 0.1192 | 0.217 | 0.597 |
| lesion side | 0.00667 | 0.78265 | 0.00728 | 0.00491 | 0.1007 | 0.0117 | 0.4844 | 0.37 | 0.5479 |
| age | 6.92E-08 | 0.22814 | 0.00817 | 1.10E-06 | 0.01438 | 0.0209 | 0.3374 | 0.254 | 0.1015 |
| model | <2.2e-16 | 0.0127 | <2.2e-16 | <2.2e-16 | 0.00175 | 7.44E-14 | 0.2523 | 0.394 | 0.2728 |

## 4 Discussion

This study evaluated the association between NIHSS score and lesion volume by vascular territory, specifically the MCA and PCA territories. We confirmed our hypothesis that right MCA ischemic strokes are larger than left MCA ischemic strokes, especially for higher NIHSS scores. We also confirmed that the NIHSS significantly depends on the infarct side of MCA strokes, after correcting for lesion volume and other covariates. Although generally in agreement with previous studies^8, 11, 12^, this study was substantially larger than previous studies (n=153-312), and evaluated the bias for separate vascular territories. We also controlled for age and sex, as both variables may be deferentially associated with infarct volume.

Previous authors have explained the hemispheric bias as a reflection of greater points given for language deficits (typically left hemisphere symptom) than hemispatial neglect (typically thought of as right hemisphere symptom). Gottesman and colleagues found supplementing the NIHSS with more points for neglect (assessed with line cancellation and visual extinction) could correct the bias. This additive approach would require 2-3 minutes but could potentially correct the left hemisphere bias^12^. This bias could be corrected in other ways (e.g. eliminating orientation and commands), but this approach would yield a less complete neurological exam. It is important to recognize that the NIHSS does not capture right cortical dysfunction. Other right, mostly MCA, cortical functions include empathy^16, 17^, recognition and expression of affective prosody (tone of voice to convey emotions^18, 19^, recognition of facial expression^20^, awareness of deficits^21^, integration of information (getting the “big picture”)^22, 23^, understanding humor and metaphor^24, 25^, and so on. However, these cortical functions are more difficult to assess reliably at bedside.

These findings may have implications for future research protocols and clinical practices that utilize the NIHSS. Some treatment protocols (e.g., involving endovascular therapy) have excluded patients with low NIHSS scores. As such, patients with large volume right hemisphere stroke and low NIHSS scores (e.g., right temporal strokes that spare motor functions) may be under-treated. These patients may be left with disabling deficits that substantially impede social function and human relationships, such as failure to empathize, understand emotional tone of voice or facial expression or humor). Likewise, use of a “diffusion-clinical mismatch”^26–28^ that uses an NIHSS score in comparison the volume of ischemia on DWI, has been advocated for thrombectomy up to 24 hours post-onset of stroke. Patients with right inferior division MCA strokes are likely not to meet the clinical criteria for this important intervention but are likely to have disabling sequelae. Thus, the NIHSS alone may not be optimal for determining the lower limits of stroke treatment eligibility, specifically for right MCA stroke patients.

## Data Availability

The data that supports this study is available in Zenodo https://doi.org/10.5281/zenodo.5722286

## Data Availability

The data that supports this study is available in Zenodo https://doi.org/10.5281/zenodo.5722286^29^.

## Acknowledgements and Sources of Funding

This research was supported in part by the National Institute of Deaf and Communication Disorders, NIDCD, through R01 DC05375, R01 DC015466, P50 DC014664 (AEH, EV, MDS, AVF).

## Author contribution

EV collected the data and drafted the work. GK and AVF collected and analyzed the data. MS significantly reviewed the draft. AEH and AVF conceived and designed the study, interpreted the data, and drafted the work.

## Supplementary Tables

**Table 4.** Functions evaluated (points attributed) in NIHSS, and the expected dominance of such functions according to brain hemisphere (left or right).

|  | Left | Right |
| --- | --- | --- |
| 1a Level of consciousness (3) | X | X |
| 1b Level of consciousness – Questions (2) | X |  |
| 1c Level of consciousness – Commands (2) | X |  |
| 2 Best gaze (2) | X | X |
| 3 Visual (3) | X | X |
| 4 Facial palsy (3) | X | X |
| 5 Motor arm (4) | X | X |
| 6 Motor leg (4) | X | X |
| 7 Limb ataxia (2) | X | X |
| 8 Sensory (2) | X | X |
| 9 Best language (3) | X |  |
| 10 Dysarthria (2) | X | X |
| 11 Extinction and inattention (2) | X | X |

**Table 5.**
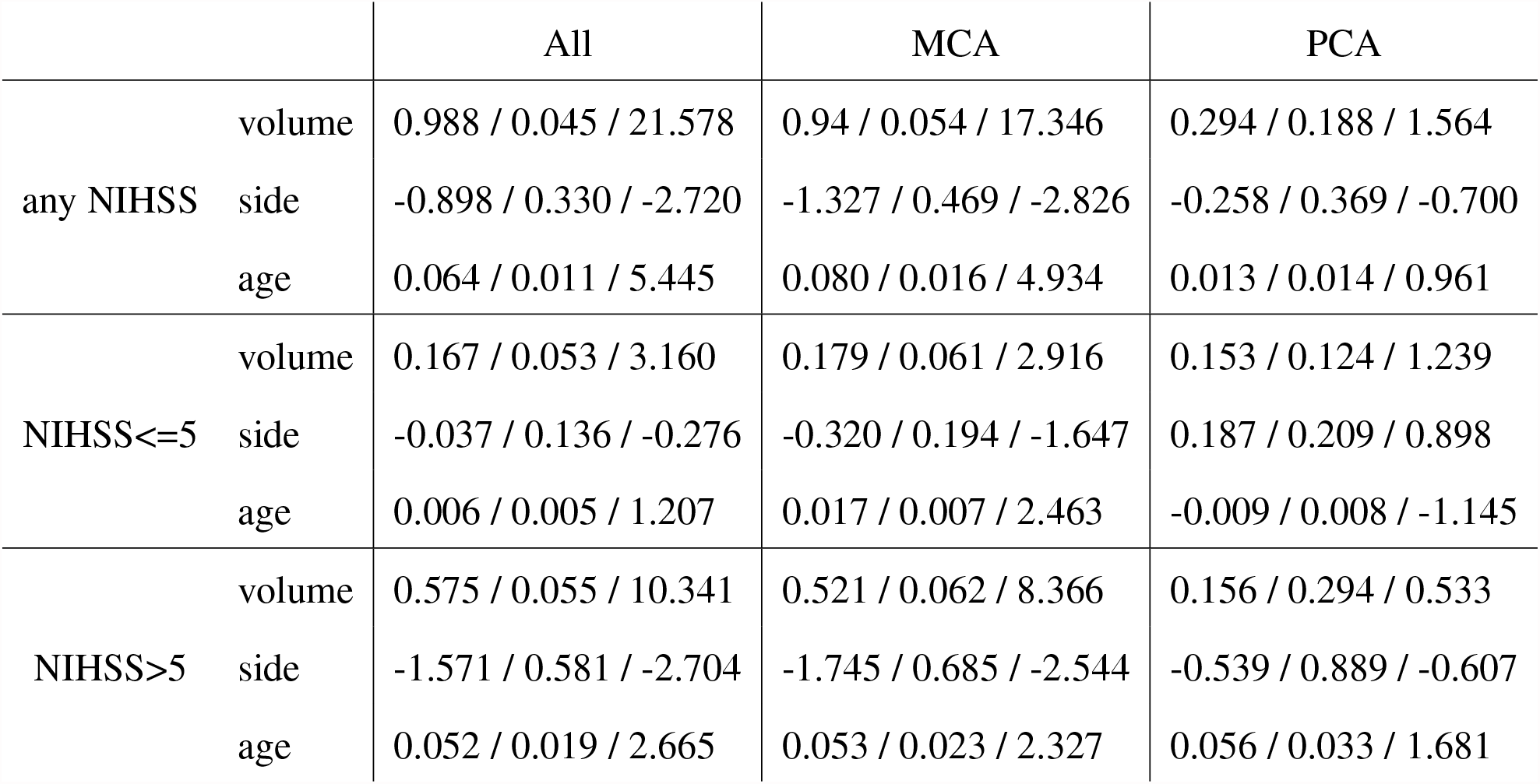
Estimates / Standard errors / t values for the covariates (lesion volume, lesion side and patient age) of the generalized linear models to predict the NIHSS.

|  |  | All | MCA | PCA |
| --- | --- | --- | --- | --- |
| any NIHSS | volume | 0.988 / 0.045 / 21.578 | 0.94 / 0.054 / 17.346 | 0.294 / 0.188 / 1.564 |
|  | side | -0.898 / 0.330 / -2.720 | -1.327 / 0.469 / -2.826 | -0.258 / 0.369 / -0.700 |
|  | age | 0.064 / 0.011 / 5.445 | 0.080 / 0.016 / 4.934 | 0.013 / 0.014 / 0.961 |
| NIHSS≤5 | volume | 0.167 / 0.053 / 3.160 | 0.179 / 0.061 / 2.916 | 0.153 / 0.124 / 1.239 |
|  | side | -0.037 / 0.136 / -0.276 | -0.320 / 0.194 / -1.647 | 0.187 / 0.209 / 0.898 |
|  | age | 0.006 / 0.005 / 1.207 | 0.017 / 0.007 / 2.463 | -0.009 / 0.008 / -1.145 |
| NIHSS>5 | volume | 0.575 / 0.055 / 10.341 | 0.521 / 0.062 / 8.366 | 0.156 / 0.294 / 0.533 |
|  | side | -1.571 / 0.581 / -2.704 | -1.745 / 0.685 / -2.544 | -0.539 / 0.889 / -0.607 |
|  | age | 0.052 / 0.019 / 2.665 | 0.053 / 0.023 / 2.327 | 0.056 / 0.033 / 1.681 |

## References

1. Brott, T. et al. Measurements of acute cerebral infarction: a clinical examination scale. Stroke 20, 864–870 (1989).

2. Goldstein, L. B., Bertels, C. & Davis, J. N. Interrater reliability of the nih stroke scale. Arch. neurology 46, 660–662 (1989).

3. Lyden, P. et al. Improved reliability of the nih stroke scale using video training. ninds tpa stroke study group. Stroke 25, 2220–2226 (1994).

4. Kasner, S. E. Clinical interpretation and use of stroke scales. The Lancet Neurol. 5, 603–612 (2006).

5. Adams, H. P. et al. Baseline nih stroke scale score strongly predicts outcome after stroke: a report of the trial of org 10172 in acute stroke treatment (toast). Neurology 53, 126–126 (1999).

6. Frankel, M. R. et al. Predicting prognosis after stroke: a placebo group analysis from the national institute of neurological disorders and stroke rt-pa stroke trial. Neurology 55, 952–959 (2000).

7. Weimar, C., Konig, I., Kraywinkel, K., Ziegler, A. & Diener, H. Age and national institutes of health stroke scale score within 6 hours after onset are accurate predictors of outcome after cerebral ischemia: development and external validation of prognostic models. Stroke 35, 158–162 (2004).

8. Fink, J. N. et al. Is the association of national institutes of health stroke scale scores and acute magnetic resonance imaging stroke volume equal for patients with right-and left-hemisphere ischemic stroke? Stroke 33, 954–958 (2002).

9. Nakajima, M. et al. Relationships between angiographic findings and national institutes of health stroke scale score in cases of hyperacute carotid ischemic stroke. Am. J. Neuroradiol. 25, 238–241 (2004).

10. Fischer, U. et al. Nihss score and arteriographic findings in acute ischemic stroke. Stroke 36, 2121–2125 (2005).

11. Woo, D. et al. Does the national institutes of health stroke scale favor left hemisphere strokes? Stroke 30, 2355–2359 (1999).

12. Gottesman, R. F. et al. The nihss-plus: improving cognitive assessment with the nihss. Behav. neurology 22, 11–15 (2009).

13. Tippett, D. C. & Hillis, A. E. Vascular aphasia syndromes. In Neurobiology of Language, 913–922 (Elsevier, 2016).

14. Kleinman, J. T. et al. Right hemispatial neglect: frequency and characterization following acute left hemisphere stroke. Brain cognition 64, 50–59 (2007).

15. Liu, C.-F. et al. Deep learning-based detection and segmentation of diffusion abnormalities in acute ischemic stroke. Commun. Medicine 1, 1–18 (2021).

16. Hillis, A. E. Inability to empathize: brain lesions that disrupt sharing and understanding another’s emotions. Brain 137, 981–997 (2014).

17. Leigh, R. et al. Acute lesions that impair affective empathy. Brain 136, 2539–2549 (2013).

18. Ross, E. D. & Monnot, M. Neurology of affective prosody and its functional–anatomic organization in right hemisphere. Brain language 104, 51–74 (2008).

19. Durfee, A. Z., Sheppard, S. M., Blake, M. L. & Hillis, A. E. Lesion loci of impaired affective prosody: A systematic review of evidence from stroke. Brain Cogn. 152, 105759 (2021).

20. Blonder, L. X. et al. Affective facial and lexical expression in aprosodic versus aphasic stroke patients. J. Int. Neuropsychol. Soc. 11, 677–685 (2005).

21. Moro, V. et al. Motor versus body awareness: voxel-based lesion analysis in anosognosia for hemiplegia and somatoparaphrenia following right hemisphere stroke. Cortex 83, 62–77 (2016).

22. Minga, J., Johnson, M., Blake, M. L., Fromm, D. & MacWhinney, B. Making sense of right hemisphere discourse using rhdbank. Top. Lang. Disord. 41, 99–122 (2021).

23. Hillis Trupe, E. & Hillis, A. Paucity vs. verbosity: Another analysis of right hemisphere communication deficits. Clin. aphasiology 15, 83–96 (1985).

24. Winner, E., Brownell, H., Happé, F., Blum, A. & Pincus, D. Distinguishing lies from jokes: Theory of mind deficits and discourse interpretation in right hemisphere brain-damaged patients. Brain language 62, 89–106 (1998).

25. Brownell, H. & Martino, G. Deficits in inference and social cognition: The effects of right hemisphere brain damage. Right hemisphere language comprehension: Perspectives from cognitive neuroscience 309 (1998).

26. Nogueira, R. G. et al. Thrombectomy 6 to 24 hours after stroke with a mismatch between deficit and infarct. New Engl. J. Medicine 378, 11–21 (2018).

27. Prosser, J. et al. Clinical-diffusion mismatch predicts the putative penumbra with high specificity. Stroke 36, 1700–1704 (2005).

28. Reineck, L. A., Agarwal, S. & Hillis, A. E. “diffusion-clinical mismatch” is associated with potential for early recovery of aphasia. Neurology 64, 828–833 (2005).

29. Faria, A. V. Nihss_802_stroke, DOI: 10.5281/zenodo.5722286 (2021).

